# Prognostic and theragnostic biomarkers in ovarian clear cell carcinoma

**DOI:** 10.1101/2020.08.21.20178830

**Authors:** Katharina Wiedemeyer, Linyuan Wang, Eun Young Kang, Shuhong Liu, Young Ou, Linda E. Kelemen, Lukas Feil, Michael S. Anglesio, Sarah Glaze, Prafull Ghatage, Gregg S. Nelson, Martin Köbel

**Author notes:** **Corresponding author:** Martin Köbel, MD, Department of Pathology and Laboratory Medicine, University of Calgary, Calgary, Alberta, Canada.

## Abstract

In this study, we aimed to test whether prognostic biomarkers can achieve a clinically relevant stratification of patients with stage I ovarian clear cell carcinoma (OCCC) and to survey the expression of 10 selected actionable targets (theragnostic biomarkers) in stage II to IV cases. From the population-based Alberta Ovarian Tumor Type study, 160 samples of OCCC were evaluated by immunohistochemistry and/or silver-enhanced in-situ hybridization for the status of 5 prognostic (p53, p16, IGF2BP3, CCNE1, FOLR1) and 10 theragnostic biomarkers (ALK, BRAF, ERBB2, ER, MET, MMR, PR, ROS1, NTRK1-3, VEGFR2). Kaplan-Meier survival analyses were performed. Cases with abnormal p53 or combined p16/IFG2BP3 abnormal expression identified a small subset of patients (6/54 cases) with stage I OCCC with an aggressive course (5-year ovarian cancer specific survival of 33.3%, compared to 91.5% in the other stage I cases). Among theragnostic targets, *ERBB2* amplification was present in 11/158 (7%) of OCCC, while MET was ubiquitously expressed in OCCC similar to a variety of normal control tissues. ER/PR showed a low prevalence of expression. No abnormal expression was detected for any of the other targets. We propose a combination of 3 biomarkers (p53, p16, IGF2BP3) to predict prognosis and the potential need for adjuvant therapy for patients with stage I OCCC. This finding requires replication in larger cohorts. Additionally, OCCC could be tested for *ERBB2* amplification for inclusion in gynecological basket trials targeting this alteration.

## Introduction

A diagnosis of ovarian clear cell carcinoma (OCCC) often represents a management challenge for oncologists as no adjuvant therapy regimen is clearly beneficial (1). OCCC is resistant to platinum-based chemotherapy, while radiation has demonstrated benefit only for localized disease (2). Thus, in the setting of localized disease, the threshold for giving adjuvant therapy might be higher for OCCC compared to other histotypes of ovarian carcinoma such as tubo-ovarian high-grade serous carcinoma for which effective primary therapy exists (3). Likewise, there is a clinical need to refine prognostic markers in OCCC to identify low risk patients, for whom ineffective adjuvant chemotherapy can be spared (4). Recently, five prognostic biomarkers (FOLR1, p16, CCNE1, p53, IGF2BP3) have been proposed for OCCC but none have yet shown proven value to refine the treatment threshold thus far (5-9).

Furthermore, there is an urgent need to identify effective treatments for OCCC patients with metastatic disease given frequent progression despite platinum-based chemotherapy. The current trend of precision oncology is based on the idea of matching genomic alterations with corresponding targeted molecular therapies (10). There have been several success stories that have resulted in remarkable improvements in cancer patient survival, such as targeted inhibitors of *ALK1* and *ROS1* in lung adenocarcinoma (11,12). Additionally, many of the actionable biomarkers identified to date, including *ALK1* and *ROS1*, can now be readily assessed by immunohistochemistry (IHC), silver-enhanced in situ hybridization, and/or chromogen in situ hybridization (CISH) on tumour tissue as cost effective and accurate surrogates for selected molecular alterations (13,14). However, the clinical utility of precision medicine is hindered by the small percentage of tumors that demonstrate targetable or so-called “theragnostic” alterations. When shotgun genomics were applied to more than 10,000 tumor samples from patients with a variety of metastatic cancers, only 11% of patients were subsequently enrolled in genomically matched clinical trials (15). Genomically driven “basket trials” combining cancers from different sites such as the NCI-MATCH study are currently underway. However, among the most common molecular alterations found in OCCC *(ARID1A, PIK3CA and TERT* promoter mutations (16)), only *ARID1A* is currently being studied in a clinical trial recruiting patients with *ARID1A*-mutated cancers for BRD4/BET inhibition (NCT03297424).

Some of the initial actionable alterations discovered in one organ have shown to be similarly actionable targets in another organ. For example, inhibitors of genetic alteration in lung carcinomas were also effective in treating colorectal adenocarcinomas with these mutations (17). Similarly, the Food and Drug Administration (FDA) approved immune therapy as the first targeted therapy for mismatch repair deficient (MMRd) neoplasms irrespective of cancer site and type (18). Clear cell renal cell carcinoma and OCCC share phenotypical similarities (clear cytoplasm) and a shared active hypoxemia pathway (19,20), providing rationale for crossover trials from renal cell cancer targeting VEGFR and MET (21-23). However, large-scale biomarker studies have not been performed in OCCC.

Therefore, the objective of the current study was 2-fold. First, we aimed to refine prognostic predictions within stage I OCCC patients using combinations of 5 previously published prognostic biomarkers. Second, we aimed to evaluate the prevalence of 10 selected therapeutically actionable targets (theragnostic biomarkers) clinically used in other cancer sites: VEGFR2, AURKA and MET (established targets in clear cell renal cell carcinoma); ALK and ROS1 (established targets in lung adenocarcinoma); ER/PR, ERRB2 (established targets in breast cancer); BRAF (established target in melanoma); MMR and NTRK (with no specific site association).

## Methods

### Study cohort and patient characteristics

OCCC cases from the Alberta Ovarian Tumor Type (AOVT) study, which included all OCCC diagnosed in Alberta, Canada, between 1979 and 2010, were obtained from the Alberta cancer registry. Histotype diagnosis was confirmed in a 2-step process including slide review and integration of diagnostic biomarker panels (24,25). Cases coded as mixed carcinomas were reviewed and no cases showed clearly separable histotype components but rather ambiguous features between clear cell and endometrioid carcinoma. These cases were reclassified either to OCCC or endometrioid carcinoma using an integration of morphology and Napsin A/PR IHC (26). In a recent publication we reported 4 OCCC with MMRd (27). On post study re-review, the morphological features were ambiguous between clear cell and endometrioid carcinoma and these cases were excluded from the current study. Ethical approval was granted by the Health Research Ethics Board of Alberta Cancer Committee (HREBA.CC-16-0161_REN3).

### Immunohistochemistry and silver-enhanced in situ hybridization

Previously built tissue microarrays were cut into 4 μm-thick sections (24). IHC for ALK, AURKA, CCNE1, ERBB2, ER, FOLR1, IGF2BP3, NTRK1-3, MET, MSH6, p16, p53, PMS2, PR, ROS1, and VEGFR2 was performed on a DAKO Omnis platform, BRAF V600E on a Leica Bond, and ERBB2 SISH on a Ventana platform. All assays derived high quality staining with high signal to noise ration. Only AURKA (clone EP1008Y) did not yield sufficient quality staining and was excluded from the study. Scoring for IGF2BP3, p16 and MMRd, were adapted from previous studies (7,8,27). Antibodies, immunohistochemical protocols, positive/negative controls and interpretation are shown in supplementary Table S1 (5,6,28–30). For the interpretation of ERBB2 IHC, we applied the 2018 American Society of Clinical Oncology/College of American Pathologists (ASCO/CAP) interpretation guidelines for breast cancer (28), defined as IHC3+: circumferential membranous staining that is complete, intense, and in >10% of tumor cells; IHC2+: weak to moderate complete membranous staining observed in >10% of tumor cells; IHC1+: incomplete membranous staining that is faint and in >10% of tumor cells; and IHC0: no staining or membranous staining that is incomplete and is faint/barely perceptible and in ≤10% of tumor cells. Some OCCC showing a single layer of tumor cells in the context of papillary or tubulocystic architectures lacked apical membranous staining. This “U-shaped” or “basolateral” staining was considered IHC2+ even if intense staining was seen. For silver-enhanced in-situ hybridization (SISH), a nuclear ERBB2 signal of ≥4.0/nucleus or an ERBB2:CEP17 ratio of ≥ 2.0 was considered amplified. MET was scored using the H score, which multiplies the distribution percentage with a 3-tier intensity to derive a maximal value of 300. Several cut-offs were tested against the primary survival endpoint.

### Statistics

Continuous variables were compared using one-way analysis of variance and categorical variables were analyzed using Pearson’s chi-squared test. Raw p-values without correction for multiple testing were reported. Univariable survival was analyzed using the Kaplan-Meier estimator with the log-rank test. The primary endpoint for survival analysis was ovarian cancer specific death, which is death due to ovarian cancer censoring death from another cause. Observations were censored at the time of last follow-up or at ten years of follow up. Analyses were performed with JMP14.0.0 (SAS Institute, North Carolina, USA).

## Results

### Stratification of cohort based on stage

We analyzed 160 OCCC from the AOVT cohort. The stage distribution was as follows: 54 (33.7%) stage I (including 18 cases stage IA), 62 (38.8%) stage II, 36 cases (22.5%) stage III and 8 cases with unknown stage information. The ovarian cancer-specific survival (OCSS) across stage was significantly different (p<0.0001, Figure 1A). The estimated 5-year OCSS for all cases was 58.5% (stage I: 85%, stage II: 66%, stage III: 31.5%). Surprisingly, the 5-year OCSS for the 18 stage IA cases was 82.5%, which was worse than the survival for all stage I OCCC patients combined (85%). Furthermore, in a stratified analysis of locally advanced disease (stages II, III and cases with an unknown stage), residual tumor was present in 31/106 (29%) of cases and associated with worse survival (p<0.0001, Figure 1B). Stage II cases without residual tumor had a 5-year OCSS of only 70.5% versus 85% for stage I cases without residual tumor. Based on this, we decided to separate the cohort into stage I and locally advanced disease (stage II, III and unknown stage) for subsequent evaluation of prognostic and theragnostic markers (Table 1). In multivariable analysis including age, stage and residual tumor, the presence of any residual tumor (HR=3.46, 95% CI 2.03-5.84, p<0.0001) and stage II or higher or unknown stage (HR=3.19, 95% CI1.63-6.81, p-0.0004) were associated with a significantly reduced ovarian cancer specific survival, while age was not (p=0.16).

**Table 1.**
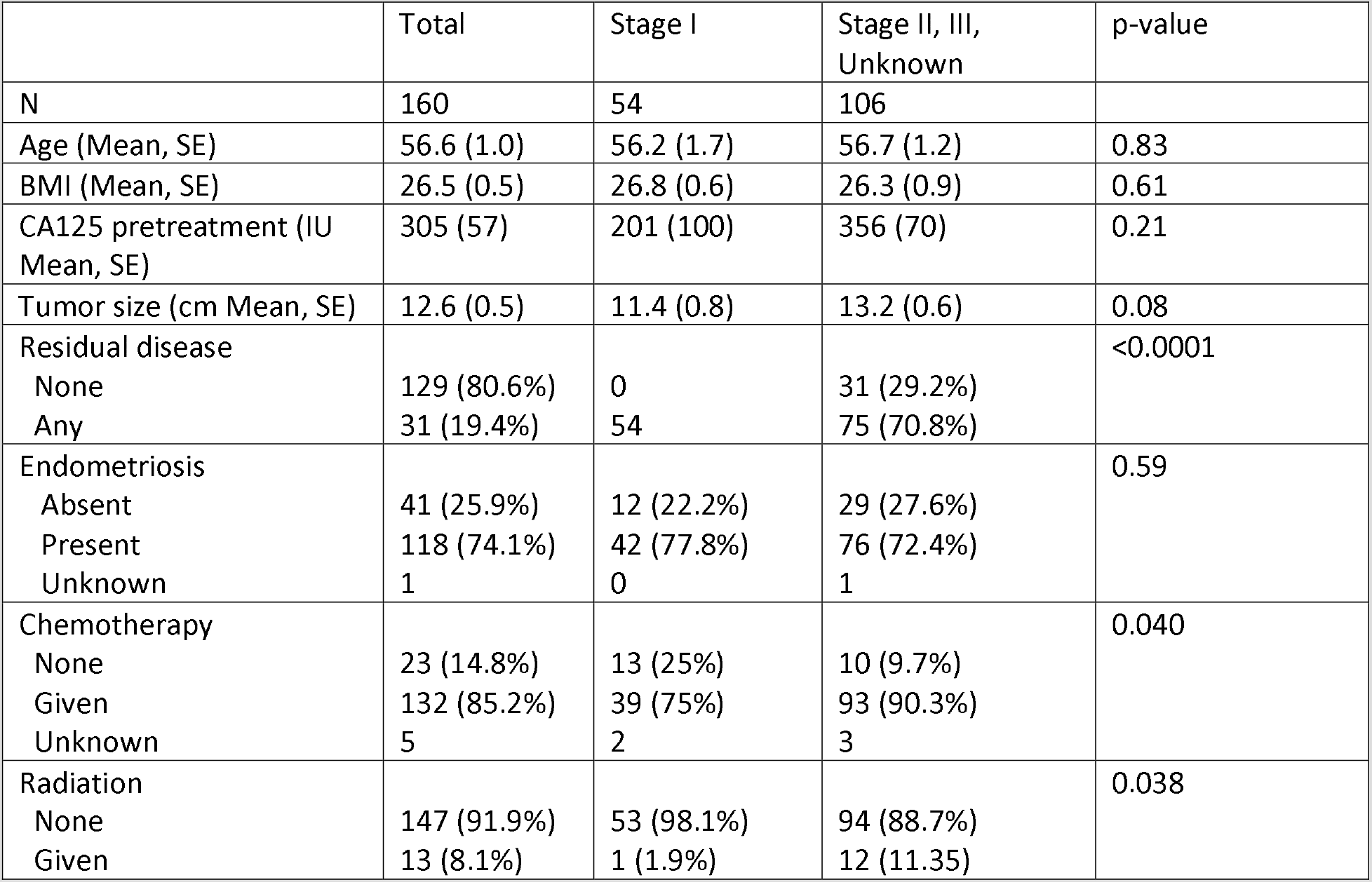
Cohort characteristics

|  | Total | Stage I | Stage II, III,<br>Unknown | p-value |
| --- | --- | --- | --- | --- |
| N | 160 | 54 | 106 |  |
| Age (Mean, SE) | 56.6 (1.0) | 56.2 (1.7) | 56.7 (1.2) | 0.83 |
| BMI (Mean, SE) | 26.5 (0.5) | 26.8 (0.6) | 26.3 (0.9) | 0.61 |
| CA125 pretreatment (IU<br>Mean, SE) | 305 (57) | 201 (100) | 356 (70) | 0.21 |
| Tumor size (cm Mean, SE) | 12.6 (0.5) | 11.4 (0.8) | 13.2 (0.6) | 0.08 |
| Residual disease |  |  |  | <0.0001 |
| None | 129 (80.6%) | 0 | 31 (29.2%) |  |
| Any | 31 (19.4%) | 54 | 75 (70.8%) |  |
| Endometriosis |  |  |  | 0.59 |
| Absent | 41 (25.9%) | 12 (22.2%) | 29 (27.6%) |  |
| Present | 118 (74.1%) | 42 (77.8%) | 76 (72.4%) |  |
| Unknown | 1 | 0 | 1 |  |
| Chemotherapy |  |  |  | 0.040 |
| None | 23 (14.8%) | 13 (25%) | 10 (9.7%) |  |
| Given | 132 (85.2%) | 39 (75%) | 93 (90.3%) |  |
| Unknown | 5 | 2 | 3 |  |
| Radiation |  |  |  | 0.038 |
| None | 147 (91.9%) | 53 (98.1%) | 94 (88.7%) |  |
| Given | 13 (8.1%) | 1 (1.9%) | 12 (11.35) |  |

**Figure 1.**
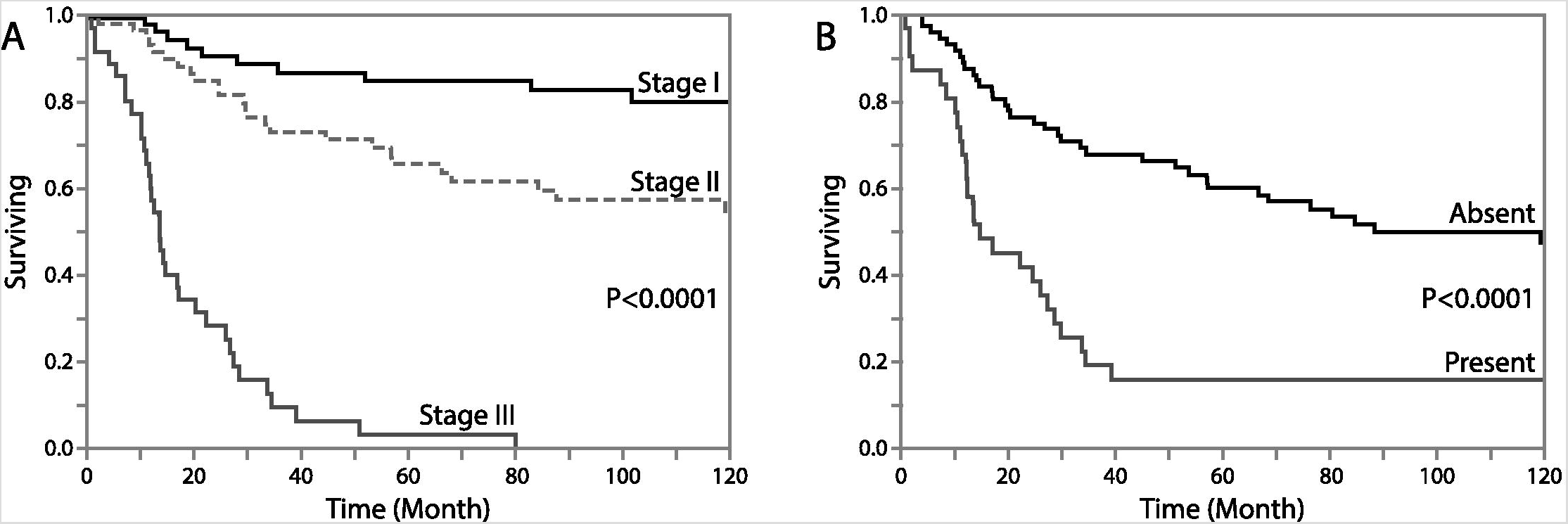
Kaplan-Meier survival curves for ovarian cancer specific survival in ovarian clear cell carcinoma (OCCC): A. all cases by stage; B. locally advanced cases stage II, III or unknown by residual disease. P-value log rank test.

### Risk stratification model for stage I ovarian clear cell carcinoma

The prevalence of the 5 selected prognostic biomarkers (p53, p16, IGF2BP3, CCNE1, FOLR1) by stage is shown in Table 2. Only p53, p16 and IGF2BP3 showed a significant association (log-rank p<0.05) with OCSS in a Kaplan-Meier analysis for stages combined, while FOLR1 (log rank p=0.21) or CCNE1 (log rank p=0.72) did not (supplementary Figure S1). We, therefore, focused on p53, p16 and IGF2BP3. Their abnormal expression was significantly associated with each other (p<0.0001, Table 3). Based on the outcome of individual combinations, we identified two risk groups. The high-risk group was characterized by p53 abnormal OCCC or combined abnormal (“double positive”) for p16 and IGF2BP3 (Figure 2A). This high-risk group of 6 out of 53 stage I cases showed a high proportion of ovarian cancer specific deaths early in the course of disease (median survival time of 20 months and 5-year OCSS of 33.3%), comparable to that of stage III cases (Figure 2B), although all 6 patients from the high-risk group received adjuvant chemotherapy in contrast to 69% from the low risk group at stage I. There were no differences in the substage distribution (pT1a/pT1c) for high versus low risk cases (p=0.99) and no significant differences in the method of primary surgery (salpingo-oophorectomy, hysterectomy, omentectomy and lymph node sampling). The 5-year OCSS of the low-risk stage I group (48/54) was 91.5% and 6.5% higher than the entire stage I group.

**Table 2.** Univariable associations with biomarker expression

|  | Total | Stage I | Stage II, III,<br>Unknown | p-value |
| --- | --- | --- | --- | --- |
| Prognostic markers (N) | 160 | 54 | 106 |  |
| p53 |  |  |  | 0.14 |
| wild type | 143 (89.4%) | 3 (5.6%) | 14 (13.2%) |  |
| mutant | 17 (10.6%) | 51 (94.4%) | 92 (86.8%) |  |
| P16/CDKN2A |  |  |  | 0.36 |
| Absent/heterogeneous | 130 (81.2%) | 8 (14.8%) | 22 (20.8%) |  |
| Block | 30 (18.8%) | 46 (85.2%) | 84 (79.2%) |  |
| IGF2BP3 |  |  |  | 0.11 |
| Absent/heterogeneous | 110 (69.6%) | 42 (77.8%) | 68 (65.4%) |  |
| Block | 48 (30.4%) | 12 (22.2%) | 35 (34.6%) |  |
| Missing | 2 | 0 | 2 |  |
| CCNE1 |  |  |  | 0.30 |
| Low | 64 (40.8%) | 19 (35.2%) | 45 (43.7%) |  |
| High | 93 (59.2%) | 35 (64.8%) | 48 (56.35) |  |
| Missing | 3 | 0 | 3 |  |
| FOLR1 |  |  |  | 0.32 |
| Absent | 73 (46.2%) | 22 (40.7%) | 51 (49.0%) |  |
| Present | 85 (53.8%) | 32 (59.3%) | 53 (51.0%) |  |
| Missing | 2 | 0 | 2 |  |
| Theranostic targets |  |  |  |  |
| MET |  |  |  | 0.68 |
| Low | 73 (45.9%) | 26 (48.2%) | 47 (44.8%) |  |
| High | 86 (53.1%) | 28 (51.8%) | 58 (55.2%) |  |
| Missing | 1 | 0 | 1 |  |
| ERBB2 |  |  |  | 0.25 |
| Non amplified | 147 (93.0%) | 52 (96.3%) | 95 (91.4%) |  |
| Amplified | 11 (7.0%) | 2 (3.7%) | 9 (8.6%) |  |
| Missing | 2 | 0 | 2 |  |
| ER |  |  |  | 0.84 |
| Absent | 133 (84.2%) | 45 (83.3%) | 88 (84.6%) |  |
| Present | 25 (15.8%) | 9 (16.7%) | 16 (15.4%) |  |
| Missing | 2 | 0 | 2 |  |
| PR |  |  |  | 0.14 |
| Absent | 9 (5.7%) | 5 (9.4%) | 4 (3.8%) |  |
| Present | 150 (94.3%) | 48 (90.6%) | 102 (96.2%) |  |
| Missing | 1 | 1 | 0 |  |

p-values calculated excluding missing data.

**Table 3.** Associations of biomarker expression

|  | P53 | P16 | IGF3BP3 | CCNE1 | FOLR1 | MET | ERRB2 | ER | PR |
| --- | --- | --- | --- | --- | --- | --- | --- | --- | --- |
| P53 | X | **<0.0001** | **<0.0001** | *0.0005* | *0.0071* | 0.68 | 0.067 | 0.36 | 0.28 |
| P16 | X | X | **<0.0001** | *0.0097* | *0.0002* | 0.12 | **0.020** | 0.82 | 0.14 |
| IGF2BP3 | X | X | X | 0.086 | *0.0066* | 0.18 | 0.071 | 0.27 | *0.040* |
| CCNE1 | X | X | X | X | **0.0048** | **0.035** | *0.025* | 0.91 | 0.25 |
| FOLR1 | X | X | X | X | X | 0.47 | 0.56 | 0.17 | 0.41 |
| MET | X | X | X | X | X | X | 0.99 | 0.60 | 0.14 |
| ERRB2 | X | X | X | X | X | X | X | **0.045** | 0.39 |
| ER | X | X | X | X | X | X | X | X | **0.0046** |
| PR | X | X | X | X | X | X | X | X | X |

Pearson chi square test, bold - positive association, italic – inverse association; p-value not corrected for multiple testing.

**Figure 2.**
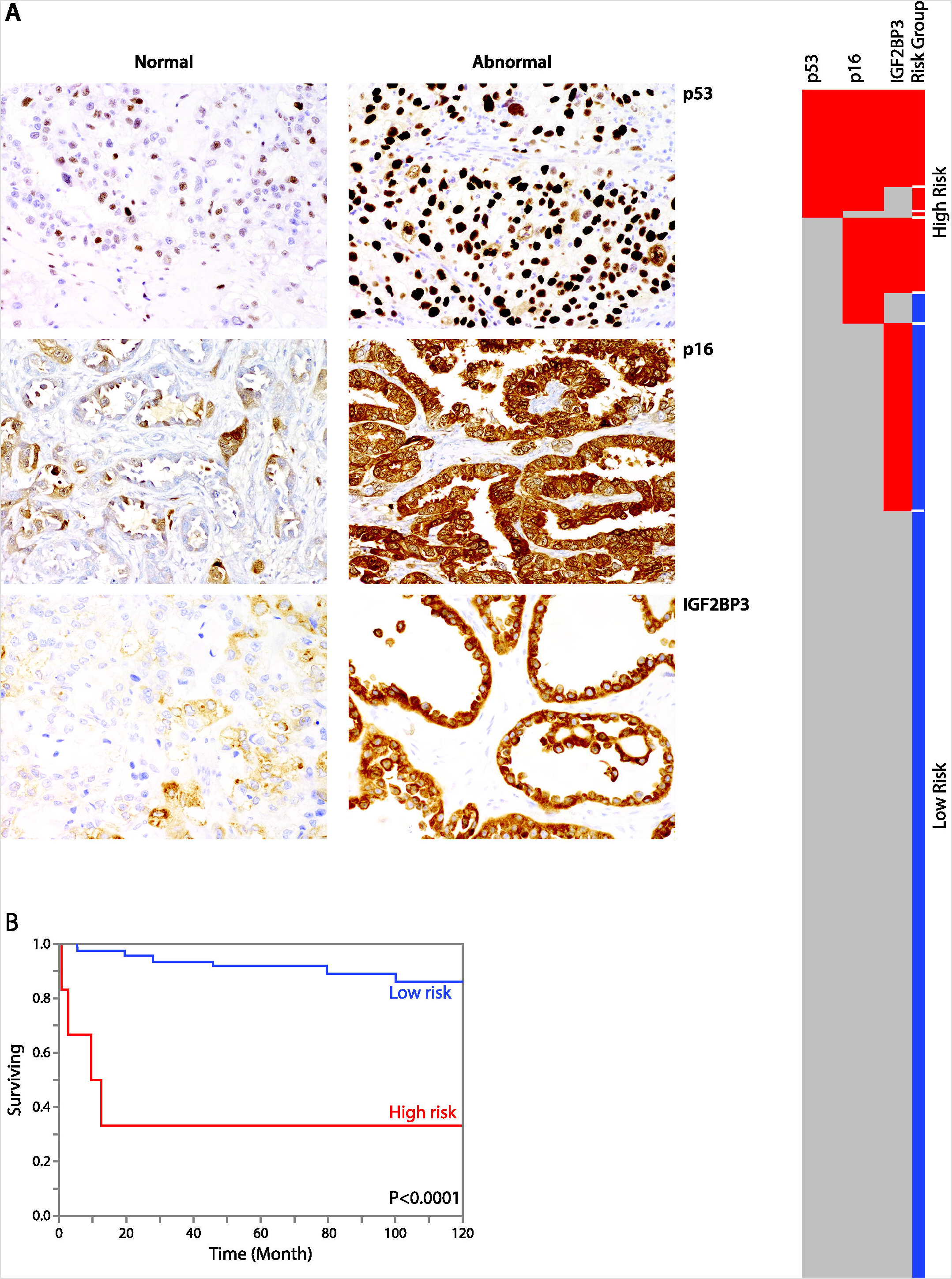
Prognostic marker model for stage I ovarian clear cell carcinoma (OCCC): A. representative of abnormal and normal expression of p53, p16 and IGF2BP3. B. Oncoplot showing 7 combinations of p53, p16 and IGF3BP3 for all 160 cases. Based on individual outcome, combination 1–4 were categorized into high-risk versus combination 5–7 into low-risk. Kaplan-Meier survival curves comparing ovarian cancer specific survival for high versus low-risk groups within stage I OCCC, p-value log rank test.

### Theragnostic targets in locally advanced ovarian clear cell carcinoma

From the 10 theragnostic targets, only ERBB2, MET, ER and PR biomarkers showed expression in OCCC (Figure 3). Their distribution by stage is shown in Table 2. For ERBB2, 18 cases (11.4%) were IHC 2+ and 4 cases (2.5%) were IHC 3+. There was excellent concordance between HER2 IHC and SISH: all IHC3+ cases were amplified by SISH, and none of the IHC0/IHC1+ cases were amplified on SISH. Among IHC2+ cases, 39% (7/18) were amplified by SISH. *ERBB2* SISH detected 11 amplified cases in total (7.0%). We performed a Kaplan-Meier survival analysis and found that the *ERBB2* amplified cases showed an unfavorable outcome across all stages (log-rank p=0.028, Figure 3). There was also a strong association of *ERBB2* amplification with p16 block-expression and abnormal p53 IHC staining (Table 4) as well as in high-risk OCCC (18.5% versus 4.6%, p=0.0096).

**Figure 3.**
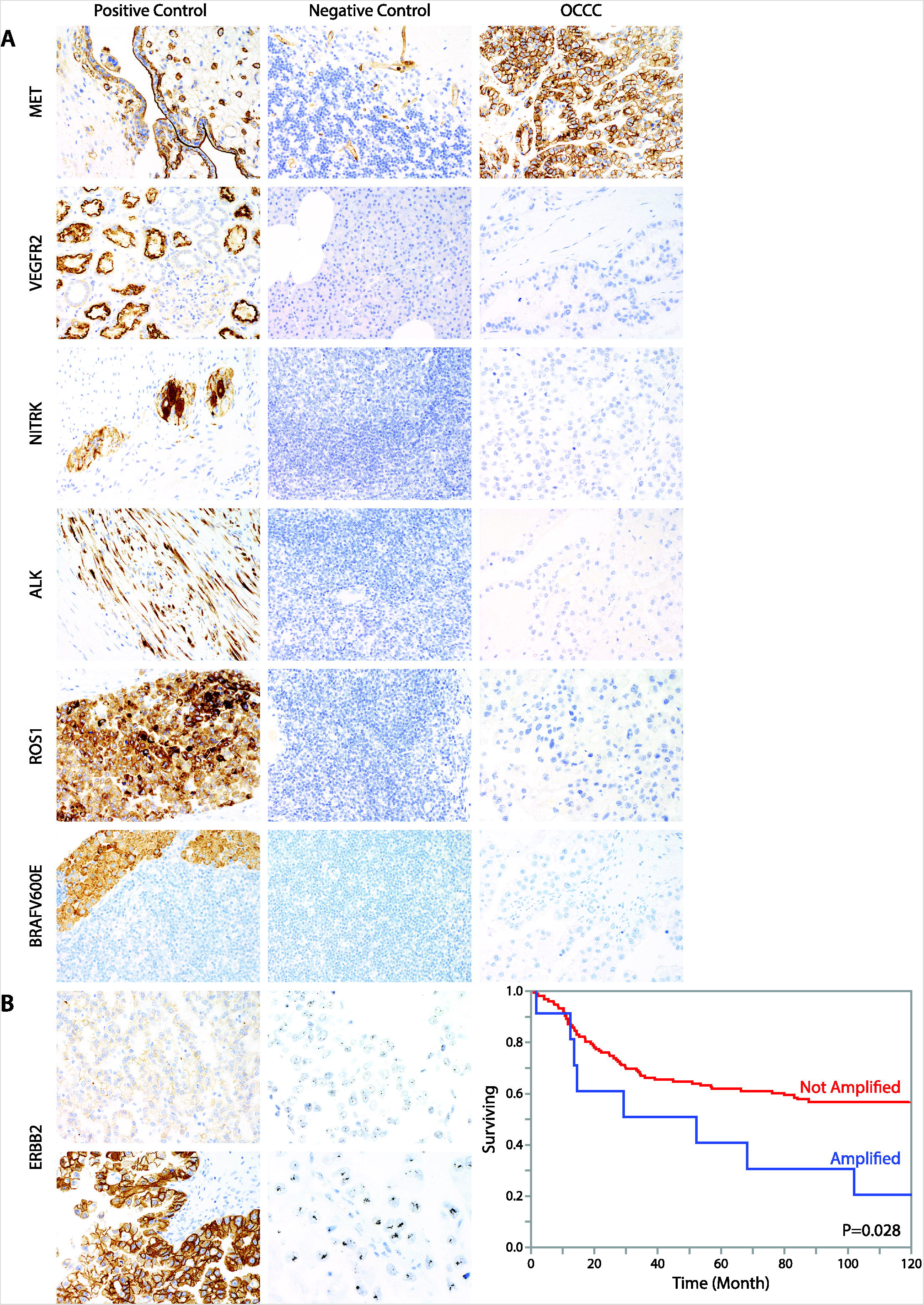
Theragnostic biomarkers in ovarian clear cell carcinoma (OCCC): A. representative images from selected theragnostic markers showing strong membranous MET staining in OCCC. The negative control brain tissue still shows MET expression in endothelial cells. VEGFR2, NTRK1–3, ALK, ROS1 and BRAF V600E were complete absent in OCCC, while strong signals were obtained from the positive controls; B. ERBB2 IHC2+ without amplification and IHC3+ with amplification by chromogenic in situ hybridization. Kaplan-Meier survival curves comparing ovarian cancer specific survival for ERBB2 amplified OCCC versus not amplified cases, P-value log rank test.

MET was highly expressed in OCCC; 97% of cases showed expression and the mean H-score was 216, SD 97. At a cut-off of H-score >200, slightly more than half of OCCC were high expressors (Table 3) MET expression was not associated with survival (several cut-offs were tested e.g. H-score >200, log rank p=0.88, Supplementary Figure S1). We observed strong MET expression in a variety of tested normal tissues including the placenta, fallopian tube, endometrium, tonsil, pancreas, liver (canalicular pattern), lung, distal renal tubules, small and large bowel (epithelium and stroma of lamina propria), and inflammatory cells including macrophages (Supplementary Figure 2). Weak MET expression was seen in squamous epithelium, thyroid and testis. From the normal tissues tested, only brain tissue was negative, although expression in endothelial cells was present.

Amongst normal tissues, VEGFR2 was expressed in the brush border of proximal renal tubules and in inflammatory cells including plasma cells; but it was absent in pancreas, liver, tonsil, and brain. Low level expression was seen in the testis. None of the OCCC showed VEGFR2 expression in the tumor epithelium nor in the microenvironment including endothelial cells. Similarly, ALK1 (0/158), BRAF (0/157), NTRK (0/158), and ROS1 (0/158) were not expressed in any of the evaluated cases. All OCCC were MMR proficient (160/160).

## Discussion

Our survey of 160 well-characterized OCCC highlights ERBB2 as a potential therapeutic target that may be exploited by off-label use of FDA approved drugs in a small subset of patients. We were also able to identify a small subgroup of stage I OCCC cases with a very aggressive biology using a combination of p53, p16, and IGF2BP3.

The search for novel targeted therapies in OCCC has been challenging. The initial clinical trial with trastuzumab, a first generation HER2 inhibitor, in ovarian carcinomas showed a dismal response of 7% in all comers of ovarian carcinomas with ERBB2 expression scores of IHC2+ or 3+ (31). One possible explanation of the low response in this trial may be a lack of powered histotype-specific analysis and lack of an accurate predictive test. As we show in our study, OCCC cases with IHC2+ were near-evenly split with only 7/18 having detectable ERBB2 amplification by SISH. There are now several high quality assays for ERBB2 IHC and SISH and criteria for interpretation have been established for breast and gastric cancers (28,32). However, consistent scoring criteria for gynecologic malignancies has not been agreed upon (33,34), Furthermore, lower level copy number gains in *ERBB2*, which have been reported to be as high as 33%, may account for some variability in IHC 2+ scores vs SISH amplification cut-offs (35). We confirm previous observations that adherence to breast cancer ASCO/CAP IHC interpretation criteria results in high concordance of IHC 0, 1+ and 3+ with the amplification status, particularly if basolateral staining is interpreted as IHC 2+ (33,36). Our study has shown a frequency of only 7% (11/158), for *ERBB2* amplification in OCCC which is half (14%, 7/50) of what has been previously reported (37). Since we used similar assays and interpretation criteria, this difference may be due to ascertainment bias. We and others have demonstrated that ERBB2 is a relevant target in a small number of OCCC. We believe *ERBB2* amplified OCCC should be included in basket trials of other gynecologic cancers such as endometrial serous and ovarian mucinous carcinoma (38,39). Additionally, further functional research is warranted to investigate the effects of intermediate level expression of ERBB2.

Among the potential theragnostic crossovers from renal clear cell carcinoma, MET but not VEGFR2 was expressed in OCCC. There was no association between MET expression and survival, and we made the sobering observation that MET was constitutively expressed at high levels in almost all normal tissues, which may explain the high toxicity of MET inhibitors such as cabozantinib in clinical trials (40). In the absence of defined molecular alterations for MET in OCCC, the rationale for clinical trials targeting MET in OCCC solely based on overexpression is not sufficient and a clinical trial failure could have been predicted (41). The antiangiogenic drug cediranib, targeting VEGFR, is currently being tested in a clinical trial of patients diagnosed with all ovarian carcinoma histotypes (NCT02502266). Cediranib inhibits VEGFR1-3. While we have not assessed the expression of VEGFR1 and VEGFR3, the lack of VEGFR2 expression in OCCC tumor cells and their microenvironment raises concerns regarding the success of this trial. Similarly, a previous trial investigating ENMD-2076 an oral multitarget kinase inhibitor against Aurora 1 and VEGFR, did not show efficacy in OCCC (22). Overall, this highlights the importance of more careful development of predictive biomarker assays for inclusion in clinical trials.

As previously reported, we observed low frequencies of ER expression and almost no PR expression, suggesting that the hormonal axis in OCCC is inactive and hormone therapy is of little clinical utility (42,43). MMRd is often identified as a potential theragnostic target in OCCC. No MMRd OCCC cases were included in the current cohort. In our previous study using the same cohort we reported 4 MMRd OCCC cases (27). Upon review, however, these cases exhibited an ambiguous phenotype with endometrioid features. We recently showed that endometrial carcinomas exhibiting “mixed” endometrioid and clear cell features are often MMRd (44). We now generally consider MMRd an endometrioid-specific alteration (27). Several recent studies reported a very low frequency of MMRd frequency in OCCC suggesting that universal MMR testing may not be justified for OCCC (9,27,45,46). However, this is in contrast to the recently published ASCO guidelines recommending MMR testing for OCCC (47). Further studies including independent biomarkers are needed to refine the diagnostic threshold between OCCC and ovarian endometrioid carcinoma for the small portion of phenotypically overlapping cases to determine whether MMRd occurs in prototypical OCCC.

IHC has excellent sensitivity and specificity for some targets (ALK1, ROS) but may suffer from limited sensitivities for certain translocations (e.g. NTRK) (13). Future studies using next generation sequencing may find uncommon alterations, but the unknown benefits of molecular drugs in the context of OCCC may prevent large scale implementation. Therefore, OCCC patients should at least be informed about the low probability of finding currently targetable alteration (16,48). More generally, our results suggest that many common theragnostic alterations are largely absent in OCCC and raise the bar to pursue different precision medicine strategies for OCCC (49,50).

Finally, we found that abnormal p53 staining or the combination of block-staining for both IGF2BP3 and p16 identifies cases of stage I OCCC with poor outcome. This high-risk subset showed a 5-year OCCC-specific survival of only 33%, compared to 91.5% in the other stage I OCCC cases. *TP53* mutations occurring in about 10% of OCCC have been associated with unfavorable outcome but do not seem to predict response to standard chemotherapy (9,46). Neither p16 nor IGF2BP3 alone appear to confer a significantly increased risk of progression at stage I. While low stage OCCC have a generally favorable outcome, validation of our results and subsequent promotion of the use of these biomarkers may be critical in identifying patients for which adjuvant therapy with radiation, rapid placement on novel management trials, or at the very least increased post-surgical surveillance, is warranted (2,3). Notably, we were not able to validate the prognostic association of CCNE1 expression or FORL1 expression as previously reported (5,6).

There are a number of limitations of the current study. The limited sample size did not allow for a study design where the cohort could be separated into hypothesis generating and validation sets. The numbers of patients in the high risk group after stratification for stage was small; hence, our findings require independent validation in a consortium-type study. Finally, owing to the tissue microarray resource, only targets assessable by IHC or CISH were evaluated.

In conclusion OCCC remains an “enigmatic disease (51)” that requires specific answers. Shot gun genomics appear to have had little impact or utility in this tumor type, suggesting that epigenomic characterization, better model systems, and generally more creative solutions may still be very much needed. We were unable to reconcile why ERBB2 has not been tested in OCCC despite being identified as a potential target for almost a decade, and one can only speculate that clinical trial design has impeded this. We propose a combination of p53, p16 and IGF2BP3 biomarkers to predict prognosis for patients with stage I OCCC, which are in need of more aggressive treatment.

## Data Availability

On request

## Acknowledgements

We thank Thomas Kryton, BFA, digital imaging specialist for Alberta Precision Laboratory for creating the composite figures. The study was supported by APL internal research support RS19-609.M.S. Anglesio is funded through a Michael Smith Foundation for Health Research Scholar Award and the Janet D. Cottrelle Foundation Scholars program managed by the BC Cancer Foundation.

## Supplementary material

**Table S1** Immunohistochemistry and silver-enhanced in-situ hybridization (SISH)

**Figure S1** Kaplan-Meier survival analysis for selected markers, entire OCCC cohort, p-value log rank

**Figure S2** MET expression in normal tissue

